## Supplementary Information for "Spatial Navigation as a Digital Marker for Clinically Differentiating Cognitive Impairment Severity"

### Supplementary Materials

#### Supplementary Information A

##### Distribution of dementia duration in the clinical sample

Supplementary Table 1 summarises the duration of dementia for all participants diagnosed with dementia. Duration of dementia was operationalised as the time (in years) since the clinical diagnosis. Across the full sample (n = 53), the mean duration of dementia since diagnosis was 3.1 years (SD = 3.0), indicating that most individuals had received a diagnosis relatively recently, with only a small proportion displaying long disease duration.

**Supplementary Table 1 | Distribution of dementia duration in the clinical sample.** Years since diagnosis of dementia among participants classified with dementia, presented by duration category.

| **Dementia duration** | **n** | **mean** | **median** | **sd** |
| --- | --- | --- | --- | --- |
| 0–1 year | 17 | 0.8 | 1.0 | 0.4 |
| 2–4 years | 28 | 2.8 | 2.0 | 0.9 |
| 5–9 years | 6 | 7.5 | 8.0 | 1.6 |
| ≥10 years | 2 | 13.5 | 13.5 | 0.7 |
| Overall | 53 | 3.1 | 2.0 | 3.0 |

#### Supplementary Information B

##### B.1: Between-group differences in across CDR stages for each SPACE task

##### We performed robust ANOVAs to examine differences in SPACE task performance across Clinical Dementia Rating (CDR) stages. Significant group effects were observed for training time (*F* = 14.7, *p* < .001, ξ = 0.657), path integration distance error (*F* = 7.0, *p* = .004, ξ = 0.587), and perspective taking error (*F* = 11.0, *p* < .001, ξ = 0.510), indicating progressive declines in performance with increasing clinical severity. No significant group differences were detected for egocentric pointing error (*F* = 2.61, *p* = .101) or mapping accuracy (*F* = 1.33, *p* = .291). Post-hoc contrasts, estimated using bootstrap-based modified one-step estimators (5,000 samples), showed that participants with higher CDR levels performed worse on training, path integration, and perspective taking, with moderate to large effect sizes (ξ up to 0.886).

**Supplementary Table 2 | Group differences in SPACE task performance by CDR scale levels.** ANOVA results and post-hoc comparisons for SPACE performance across clinical groups, with effect sizes and bootstrap confidence intervals.

| **ANOVA** | | | | | |
| --- | --- | --- | --- | --- | --- |
| **Dependent Variable** | | ***F*** | ***p*** | **Variance Explained** | **ES (**ξ**)** |
| Training time | 14.7 | | <.001 | 0.432 | 0.657 |
| Path integration error | 7 | | 0.004 | 0.345 | 0.587 |
| Pointing error | 2.61 | | 0.101 | 0.145 | 0.381 |
| Mapping R^2^ | 1.33 | | 0.291 | 0.065 | 0.256 |
| Perspective taking error | 11 | | <.001 | 0.26 | 0.51 |

| **Post-hoc comparisons** | | | | | | | |
| --- | --- | --- | --- | --- | --- | --- | --- |
|  | **CDR contrasts** | | **psi-hat** | ***p*** | **ES(ξ)** | **Lower CI** | **Upper CI** |
| Training time | 0 | 0.5 | -14.6 | 22 | 229 | -33.5 | 2.52 |
|  | 0 | 1 | -50.4 | <.001 | 717 | -85.3 | -23.28 |
|  | 0 | 2 | -102.8 | <.001 | 886 | -152.7 | -50.17 |
|  | 0.5 | 1 | -35.8 | 1 | 419 | -71.5 | -5.17 |
|  | 0.5 | 2 | -88.3 | <.001 | 761 | -140.4 | -35.55 |
|  | 1 | 2 | -52.5 | 22 | 483 | -109.3 | 7.76 |
| Path integration error | 0 | 0.5 | -7.57 | 0.44 | 55 | -31.5 | 15.9 |
|  | 0 | 1 | -89.35 | <.001 | 666 | -156.5 | -37.4 |
|  | 0.5 | 1 | -81.78 | <.001 | 714 | -146.5 | -27.8 |
| Pointing error | 0 | 0.5 | -1.67 | 0.55 | 64 | -7.87 | 4.77 |
|  | 0 | 1 | -13.23 | 0.03 | 39 | -24.82 | 1.51 |
|  | 0.5 | 1 | -11.56 | 58 | 420 | -24.33 | 3.54 |
| Mapping R2 | 0 | 0.5 | 13 | 826 | 38 | -128 | 150 |
|  | 0 | 1 | -134 | 168 | 143 | -304 | 101 |
|  | 0.5 | 1 | -147 | 151 | 250 | -333 | 98 |
| Perspective taking error | 0 | 0.5 | -1.67 | 572 | 47 | -9.76 | 5.91 |
|  | 0 | 1 | -14.9 | <.001 | 496 | -24.5 | -6.24 |
|  | 0 | 2 | -18.28 | <.001 | 661 | -32.51 | -7.43 |
|  | 0.5 | 1 | -13.22 | <.001 | 518 | -23.06 | -4.55 |
|  | 0.5 | 2 | -16.61 | <.001 | 613 | -31.07 | -5.64 |
|  | 1 | 2 | -3.38 | 476 | 135 | -17.42 | 8.85 |
| *Note: Bootstrap method, modified one-step estimator, 5000 samples, projection distances. Effect sizes: small ≈ 0.10, medium ≈ 0.30, large ≈ 0.50.* | | | | | | | |

##### B.2: Robust ANOVA excluding participants with CDR 3

Because the number of individuals with CDR 3 was small (n = 3), we repeated the ANOVAs after excluding these participants to ensure they did not disproportionately influence group effects. Here, we only included the training and perspective taking tasks in the analysis because only two participants from the moderate group completed the path integration, pointing and mapping tasks. Results revealed significant CDR group differences in training time (*F* = 11.6, *p* = .001, ξ = 0.613) and perspective taking error (*F* = 11.3, *p* < .001, ξ = 0.536).

#### Supplementary Information C

##### Logistic regression coefficients for classification models

We fitted logistic regression models to classify CDR stages using demographic variables (age, education, gender) together with the SPACE measures identified in the ANOVAs (training time, path integration distance error and perspective taking error). In contrasts involving CDR 0 vs CDR 1, both training time (β = 0.03, *p* < 0.001) and path integration distance error (β = 0.01, *p* < 0.001) were significant predictors. For CDR 0 vs CDR 2+ contrast, only training time (β = 0.03, *p* < 0.001) was a significant predictor. For the CDR 0.5 vs CDR 1 contrast, all training time ((β = 0.02, *p* = 0.018), path integration distance error (β = 0.02, *p* < 0.001), and perspective taking (β = 0.06, *p* = 0.006) were significant predictors. Finally, for the CDR 0.5 vs CDR 2+ contrast, training time (β = 0.02, *p* = 0.002) and perspective taking (β = 0.05, *p* = 0.015) were significant predictors.

##### Supplementary Table 3 | Logistic regression coefficients for classification models.

| **Model** | **Predictor** | **β** | **SE** | **Z** | **p** | **OR** |
| --- | --- | --- | --- | --- | --- | --- |
| 0 vs. 0.5 | Intercept | -3.02 | 1.84 | -1.65 | 0.099 | 0.05 |
|  | Age | 0.01 | 0.03 | 0.43 | 0.667 | 1.01 |
|  | Education [University]: |  |  |  |  |  |
|  | No formal Education | 0.15 | 0.83 | 0.18 | 0.859 | 1.16 |
|  | Elementary school | 0.56 | 0.53 | 1.05 | 0.295 | 1.74 |
|  | Middle school | 0.53 | 0.37 | 1.41 | 0.158 | 1.69 |
|  | High school | 0.61 | 0.39 | 1.56 | 0.119 | 1.84 |
|  | Gender [Male]: |  |  |  |  |  |
|  | Female | -0.11 | 0.29 | -0.38 | 0.706 | 0.90 |
|  | Training Time | 0.01 | 0.00 | 2.02 | 0.044 | 1.01 |
|  | PI Distance | 0.00 | 0.00 | -0.56 | 0.577 | 1.00 |
|  | Perspective Error | -0.01 | 0.01 | -0.70 | 0.484 | 0.99 |
| 0 vs. 1 | Intercept | -6.71 | 4.68 | -1.43 | 0.152 | 0.00 |
|  | Age | -0.12 | 0.06 | -1.85 | 0.065 | 0.89 |
|  | Education [University]: |  |  |  |  |  |
|  | No formal Education | -13.96 | 1437.72 | -0.01 | 0.992 | 0.00 |
|  | Elementary school | 3.52 | 1.36 | 2.58 | 0.01 | 33.66 |
|  | Middle school | 3.01 | 1.25 | 2.40 | 0.017 | 20.24 |
|  | High school | 2.75 | 1.27 | 2.16 | 0.031 | 15.63 |
|  | Gender [Male]: |  |  |  |  |  |
|  | Female | -1.56 | 0.70 | -2.23 | 0.026 | 0.21 |
|  | Training Time | 0.03 | 0.01 | 3.36 | <.001 | 1.03 |
|  | PI Distance | 0.01 | 0.00 | 3.36 | <.001 | 1.01 |
|  | Perspective Error | 0.03 | 0.02 | 1.45 | 0.146 | 1.03 |
| 0 vs. 2+ | Intercept | -31.84 | 2072.00 | -0.02 | 0.988 | 0.00 |
|  | Age | 0.01 | 0.07 | 0.18 | 0.858 | 1.01 |
|  | Education [University]: |  |  |  |  |  |
|  | No formal education | 17.77 | 2071.99 | 0.01 | 0.993 | 5.22E+07 |
|  | Elementary school | 18.02 | 2071.99 | 0.01 | 0.993 | 6.68E+07 |
|  | Middle school | 18.30 | 2071.99 | 0.01 | 0.993 | 8.85E+07 |
|  | High school | 16.75 | 2071.99 | 0.01 | 0.994 | 1.88E+07 |
|  | Gender [Male]: |  |  |  |  |  |
|  | Female | 0.31 | 0.91 | 0.34 | 0.736 | 1.36 |
|  | Training Time | 0.03 | 0.01 | 3.82 | <.001 | 1.03 |
|  | Perspective Error | 0.04 | 0.02 | 1.87 | 0.061 | 1.04 |
| 0.5 vs. 1 | Intercept | -6.94 | 4.59 | -1.51 | 0.131 | 0.00 |
|  | Age | -0.11 | 0.06 | -1.91 | 0.057 | 0.90 |
|  | Education [University]: |  |  |  |  |  |
|  | No formal education | -15.72 | 2112.53 | -0.01 | 0.994 | 0.00 |
|  | Elementary school | 1.01 | 1.49 | 0.67 | 0.5 | 2.74 |
|  | Middle school | 1.17 | 1.38 | 0.85 | 0.397 | 3.23 |
|  | High school | 0.40 | 1.43 | 0.28 | 0.779 | 1.49 |
|  | Gender [Male]: |  |  |  |  |  |
|  | Female | -0.76 | 0.71 | -1.08 | 0.281 | 0.47 |
|  | Training Time | 0.02 | 0.01 | 2.36 | 0.018 | 1.02 |
|  | PI Distance | 0.02 | 0.00 | 3.64 | <.001 | 1.02 |
|  | Perspective Error | 0.06 | 0.02 | 2.75 | 0.006 | 1.06 |
| 0.5 vs. 2+ | Intercept | -31.74 | 1985.57 | -0.02 | 0.987 | 0.00 |
|  | Age | 0.04 | 0.07 | 0.62 | 0.535 | 1.04 |
|  | Education [University]: |  |  |  |  |  |
|  | No formal education | 16.68 | 1985.56 | 0.01 | 0.993 | 1.76E+07 |
|  | Elementary school | 16.75 | 1985.56 | 0.01 | 0.993 | 1.87E+07 |
|  | Middle school | 16.97 | 1985.56 | 0.01 | 0.993 | 2.34E+07 |
|  | High school | 15.61 | 1985.56 | 0.01 | 0.994 | 6.04E+06 |
|  | Gender [Male]: |  |  |  |  |  |
|  | Female | 0.78 | 0.85 | 0.92 | 0.36 | 2.19 |
|  | Training Time | 0.02 | 0.01 | 3.10 | 0.002 | 1.02 |
|  | Perspective Error | 0.05 | 0.02 | 2.44 | 0.015 | 1.05 |
| 1 vs. 2+ | Intercept | -25.10 | 2250.86 | -0.01 | 0.991 | 0.00 |
|  | Age | 0.03 | 0.07 | 0.42 | 0.672 | 1.03 |
|  | Education [University]: |  |  |  |  |  |
|  | No formal education | 33.97 | 3570.68 | 0.01 | 0.992 | 5.67E+14 |
|  | Elementary school | 14.92 | 2250.86 | 0.01 | 0.995 | 3.02E+06 |
|  | Middle school | 16.04 | 2250.86 | 0.01 | 0.994 | 9.22E+06 |
|  | High school | 15.95 | 2250.86 | 0.01 | 0.994 | 8.42E+06 |
|  | Gender [Male]: |  |  |  |  |  |
|  | Female | 1.31 | 0.92 | 1.43 | 0.153 | 3.70 |
|  | Training Time | 0.01 | 0.01 | 1.66 | 0.097 | 1.01 |
|  | Perspective Error | 0.03 | 0.02 | 1.50 | 0.134 | 1.03 |

#### Supplementary Information D

##### Diagnostic Performance of SPACE and Standard Neuropsychological Tests Across CDR Levels

We compared the classification performance of SPACE with that of standard neuropsychological assessments across CDR contrasts. AUC values were computed at the Youden-optimal cutoff, and *p* values were obtained using Bonferroni-corrected DeLong’s test. For CDR 0 vs 0.5, all predictors demonstrated modest discrimination, with AUCs ranging from 0.60 to 0.73, and no measure outperformed SPACE. For CDR 0 vs 1 and CDR 0 vs 2+, SPACE demonstrated high discrimination (AUCs = 0.94 and 0.95), comparable to the MoCA, animal naming, and other executive function measures. For CDR 0.5 vs 1 and CDR 0.5 vs 2+, SPACE performance remained strong (AUC = 0.91 for both), similar to MoCA and several of the other neuropsychological assessments. For CDR 1 vs 2+, all predictors showed moderate to high performance (AUC range = 0.77–0.92). Together, these analyses indicate that SPACE performs on par with leading neuropsychological instruments across clinical contrasts.

**Supplementary Table 4 | AUC for each predictor across CDR contrasts.** AUC values reflect model performance at the Youden-optimal cutoff for discriminating between CDR levels. P-values are derived from DeLong’s test comparing each neuropsychological test to SPACE (reference model), with Bonferroni correction applied across predictors within each CDR contrast.

| **Outcome** | **SPACE** | **MoCA** | **QDRS** | **TMTA** | **TMTB** | **Maze task** | **DCT** | **Animal** | **Dual task** |
| --- | --- | --- | --- | --- | --- | --- | --- | --- | --- |
| AUC, *p*  (sensitivity, specificity) | | | | | | | | | |
| **CDR 0 vs 0.5** | 0.61, *Ref*  (0.47-0.79) | 0.73, 0.169 (0.51-0.86) | 0.69, 1.000 (0.66-0.63) | 0.65, 1.000 (0.46-0.78) | 0.65, 1.000 (0.38-0.88) | 0.60, 1.000 (0.49-0.71) | 0.62, 1.000 (0.52-0.74) | 0.66, 1.000 (0.36-0.94) | 0.60, 1.000 (0.54-0.66) |
| **CDR 0 vs 1** | 0.94, *Ref*  (1.00-0.85) | 1.00, 0.021 (1.00-0.98) | 0.80, 0.010 (0.70-0.77) | 0.87, 0.284 (0.82-0.81) | 0.94, 1.000 (0.85-0.90) | 0.81, 0.010 (0.82-0.75) | 0.87, 0.247 (0.77-0.81) | 0.96, 1.000 (0.85-0.97) | 0.79, 0.007 (0.62-0.84) |
| **CDR 0 vs 2+** | 0.95, *Ref*  (0.94-0.88) | 1.00, 0.163 (1.00-1.00) | 0.81, 0.168 (0.90-0.69) | 0.96, 1.000 (0.94-0.89) | 0.98, 1.000 (0.94-0.97) | 0.90, 1.000 (0.71-0.94) | 0.96, 1.000 (0.94-0.88) | 1.00, 0.213  (1.00-0.98) | 0.85, 0.989 (0.77-0.87) |
| **CDR 0.5 vs 1** | 0.91, *Ref*  (0.95-0.78) | 0.93, 1.000 (1.00-0.74) | 0.70, 0.003 (0.67-0.68) | 0.76, 0.025 (0.88-0.57) | 0.82, 0.405 (0.82-0.70) | 0.73, 0.008 (0.85-0.67) | 0.79, 0.098 (0.94-0.52) | 0.86, 1.000 (0.85-0.74) | 0.71, 0.004 (0.91-0.45) |
| **CDR 0.5 vs 2+** | 0.91, *Ref*  (0.88-0.85) | 0.99, 0.266 (1.00-0.96) | 0.81, 1.000 (0.90-0.65) | 0.91, 1.000 (0.94-0.75) | 0.91, 1.000 (0.81-0.88) | 0.85, 1.000 (0.71-0.89) | 0.92, 1.000 (0.82-0.88) | 0.97, 0.892 (0.94-0.94) | 0.82, 1.000 (0.69-0.85) |
| **CDR 1 vs 2+** | 0.83, *Ref*  (0.69-0.91) | 0.92, 1.000 (0.82-0.88) | 0.86, 1.000 (0.80-0.85) | 0.85, 1.000 (0.77-0.85) | 0.77, 1.000 (0.75-0.71) | 0.83, 1.000 (0.71-0.94) | 0.85, 1.000 (0.94-0.59) | 0.86, 1.000 (0.71-0.88) | 0.77, 1.000 (0.69-0.88) |

#### Supplementary Information E

##### E.1: Classification accuracy of SPACE and neuropsychological tests across clinical consensus diagnosis

We conducted AUC analyses for each diagnostic contrast, reporting sensitivity and specificity at the optimal cut-off. The *p*-values reflect statistical comparisons between each test and SPACE (as the reference model). For NCI vs Dementia, SPACE showed excellent discrimination (AUC = 0.94), with MoCA performing significantly better (AUC = 1.00, *p* = .006) and QDRS performing worse (AUC = 0.82, *p* = .037). All other tests did not differ significantly from SPACE. For NCI vs MCI, MoCA outperformed SPACE (AUC = 0.86 vs 0.72, *p* = .002), while all remaining assessments were statistically comparable. For MCI vs Dementia, no cognitive measure differed significantly from SPACE, which showed high discriminatory performance (AUC = 0.87).

**Supplementary Table 5 | Diagnostic accuracy of cognitive and digital assessments against clinical consensus diagnosis.**

| AUC, *p*  (Sensitivity, Specificity) | | | | | | | | |
| --- | --- | --- | --- | --- | --- | --- | --- | --- |
| Outcome | SPACE | MoCA | QDRS | TMTA | TMTB | Maze | DCT | Animal |
| NCI vs Dementia | 0.94, Ref (1.00, 0.85) | 1.00, 0.006 (1.00, 1.00) | 0.82, 0.037 (0.71, 0.80) | 0.92, 1.000 (0.94, 0.80) | 0.96, 1.000 (0.94, 0.88) | 0.83, 0.053 (0.82, 0.77) | 0.87, 0.593 (0.82, 0.78) | 0.99, 0.063 (1.00, 0.92) |
| NCI vs MCI | 0.72, Ref (0.60, 0.76) | 0.86, 0.002 (0.77, 0.84) | 0.74, 1.000 (0.63, 0.76) | 0.77, 0.895 (0.67, 0.77) | 0.81, 0.103 (0.62, 0.88) | 0.74, 1.000 (0.75, 0.65) | 0.78, 1.000 (0.68, 0.77) | 0.80, 0.223 (0.65, 0.81) |
| MCI vs Dementia | 0.87, Ref (0.94, 0.73) | 0.91, 1.000 (1.00, 0.72) | 0.71, 0.214 (0.71, 0.67) | 0.73, 0.411 (0.71, 0.78) | 0.76, 1.000 (0.82, 0.65) | 0.72, 0.318 (0.65, 0.75) | 0.73, 0.450 (0.76, 0.70) | 0.90, 1.000 (0.88, 0.75) |

##### E.2: MoCA regression

To evaluate convergent validity between SPACE and the MoCA (the best-performing assessment), we conducted a hierarchical regression analysis (Supplementary Table 6). In the first step, we entered demographic variables (age, gender, and education), which accounted for 19.1% of the variance in MoCA scores (*F*_(6,247)_ = 9.74, *p* < .001). In the second step, we added all SPACE task measures as predictors. This substantially improved model fit, explaining 50.5% of the total variance in MoCA (*F*_(12,241)_ = 20.50, *p* < .001), corresponding to a significant increase in explained variance (ΔR² = 0.314, *p* < .001). All SPACE tasks emerged as significant predictors of MoCA scores, even after controlling for demographic factors.

**Supplementary Table 6 | Model fit and comparison between hierarchical regression models to predict MoCA.**

| Model Fit Measures | | | | | | |
| --- | --- | --- | --- | --- | --- | --- |
|  | | | **Overall Model Test** | | | |
| **Model** | **R** | **R²** | **F** | **df1** | **df2** | **p** |
| 1 | 0.437 | 0.191 | 9.74 | 6 | 247 | <.001 |
| 2 | 0.711 | 0.505 | 20.50 | 12 | 241 | <.001 |
| *Note.* Models estimated using sample size of N=254 | | | | | | |

| **Predictor** | **Estimate** | **SE** | **t** | **p** |
| --- | --- | --- | --- | --- |
| Interceptᵃ | 35.80144 | 3.38906 | 10.5638 | <.001 |
| Age | 0.03654 | 0.03517 | 1.0390 | 0.300 |
| Gender: _Female - Male_ | 0.92017 | 0.41705 | 2.2064 | 0.028 |
| Education: _Primary – No formal education_ | -3.23151 | 1.22610 | -2.6356 | 0.009 |
| Education: _Secondary – No formal education_ | -0.81176 | 1.15777 | -0.7011 | 0.484 |
| Education: _High school – No formal education_ | 0.08665 | 1.16639 | 0.0743 | 0.941 |
| Education: _University – No formal education_ | 0.96165 | 1.19181 | 0.8069 | 0.421 |
| Training Time | -0.03321 | 0.00450 | -7.3821 | <.001 |
| PI Distance | -0.00789 | 0.00235 | -3.3564 | <.001 |
| Pointing Error | -0.04404 | 0.01120 | -3.9315 | <.001 |
| Mapping R² | -1.69874 | 0.67898 | -2.5019 | 0.013 |
| Memory Correct | 0.04065 | 0.00935 | 4.3477 | <.001 |
| Perspective Error | -0.04344 | 0.01068 | -4.0662 | <.001 |

#### Supplementary Information F

##### F.1: Relationships between the path integration task and subsequent pointing and mapping tasks.

To examine whether inter-task relationships differed as a function of path integration ability, we divided participants into two groups (good and poor path integrators) based on the median path integration distance error (205 m). Within each subgroup, we computed pairwise Spearman’s rank correlations between path integration distance error and performance on the other SPACE tasks (pointing error, mapping R^2^, perspective taking error, and memory percentage correct). As shown in Supplementary Fig. 1, good path integrators exhibited a coherent pattern in which greater path integration error was associated with greater pointing error (ρ = 0.19) and lower mapping accuracy (ρ = −0.29). These relationships were not observed in poor path integrators, where correlations with pointing (ρ = 0.04) and mapping (ρ = −0.08) were small and not significant. These results suggest that participants who successfully encode landmark information during path integration also perform well in the subsequent pointing and mapping tasks. In contrast, those who perform poorly in the path integration task show decoupled performance in the pointing and mapping tasks. Notably, perspective taking error showed a modest positive association with path integration error in both groups (good: ρ = 0.23; poor: ρ = 0.20), consistent with the idea that perspective taking draws on spatial updating abilities but does not rely on prior landmark encoding.

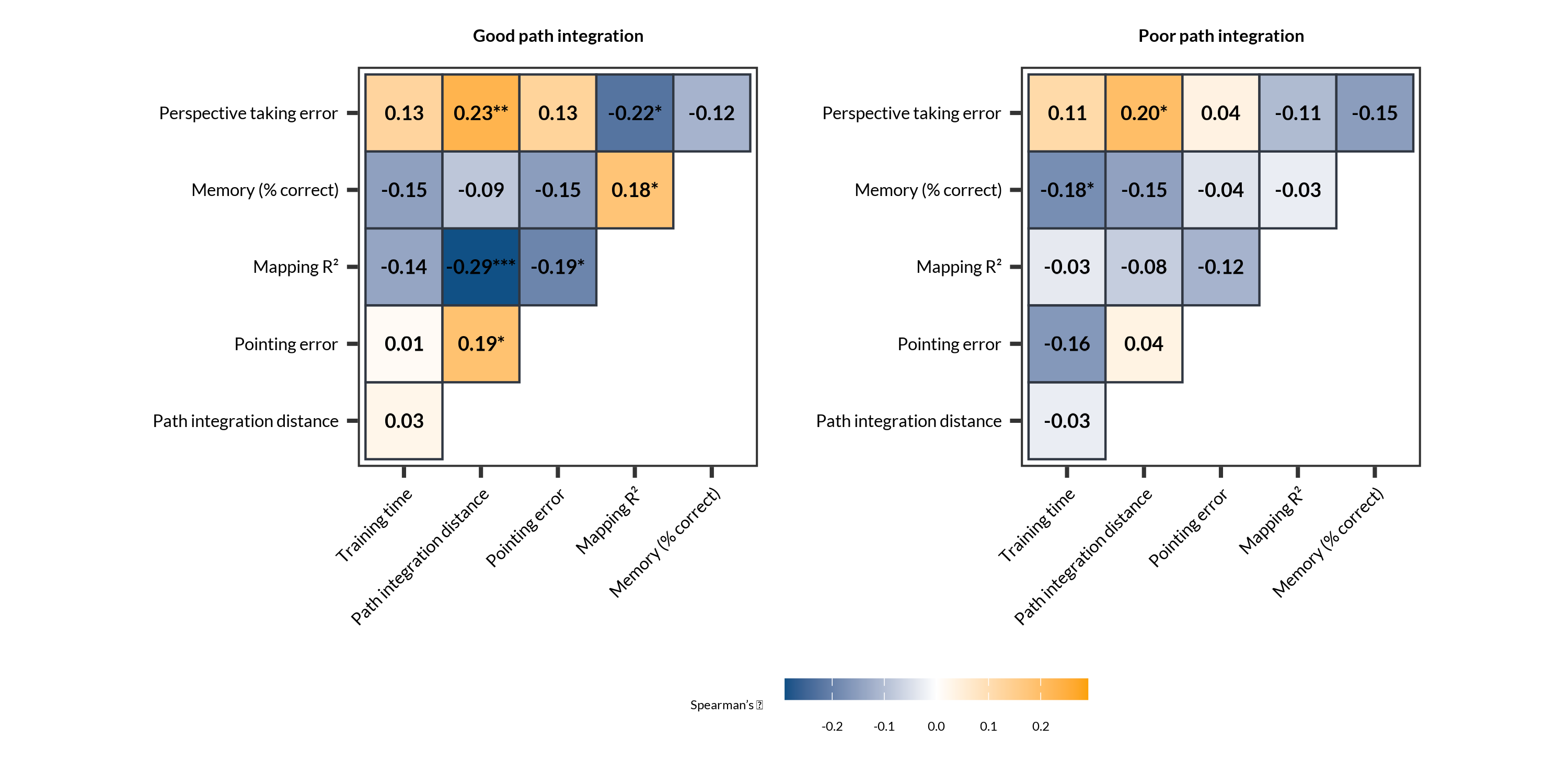
**Supplementary Figure 1 | Exploratory correlations between SPACE task performance in participants with good *vs* poor path integration ability.** Spearman’s rank correlations were computed separately for participants below (left panel) and above (right panel) the median path integration distance error (median = 205 m).

##### F.2: ****Associations between path integration and pointing and mapping task performance across CDR levels****

To clarify the relationship between the path integration, pointing and mapping tasks, we conducted additional robust regression analyses examining whether clinical status and path integration performance jointly explained variation in pointing and mapping. For pointing error, the model was significant (adjusted R² = .039). Path integration distance error was a significant predictor (β = 0.0266, SE = 0.0134, t = 1.98, *p* = 0.048), indicating that greater path integration error was associated with larger pointing error. Compared to CDR 0, participants with CDR = 1 showed significantly greater pointing error (β = 10.73, SE = 5.14, t = 2.09, *p* = 0.038), whereas the CDR 0.5 group did not differ significantly (β = 1.33, *p* = 0.60). For mapping performance (R²), the model was again significant (adjusted R² = .028). PI distance error significantly predicted mapping performance (β = −0.00066, SE = 0.00025, t = −2.67, *p* = 0.008), such that greater path integration error was associated with poorer mapping accuracy. Participants with CDR 1 exhibited significantly poorer mapping performance relative to CDR 0 (β = 0.164, SE = 0.079, t = 2.09, *p* = 0.038), while the CDR 0.5 group did not differ from CDR 0 (β = −0.007, *p* = 0.88). Across both models, higher path integration distance error consistently predicted poorer pointing and mapping performance. Impairments were most evident in individuals with CDR 1, whereas participants with CDR 0.5 showed performance comparable to CDR 0.

##### F.3: AUC analyses for SPACE including pointing and mapping tasks

Because path integration did not fully account for the observed effects in pointing and mapping, we repeated the ROC analyses to determine whether including these two measures altered SPACE’s diagnostic performance. AUC values were recalculated for all CDR group contrasts, both with and without the inclusion of pointing and mapping. As shown in Supplementary Table 7, the inclusion of pointing and mapping measures produced only minimal changes in AUC across comparisons. For CDR 0 vs 0.5, AUC decreased slightly from 0.613 to 0.603 (ΔAUC = –0.011), and for CDR 0 vs 1, AUC increased marginally from 0.945 to 0.964 (ΔAUC = 0.019). Similarly, for CDR 0.5 vs 1, AUC increased from 0.914 to 0.952 (ΔAUC = 0.038). None of these differences reached statistical significance (all *p* Bonferroni-corrected *p* ≥ 0.37). Comparisons involving CDR ≥ 2 were not evaluated due to the small number of participants in this subgroup. Overall, these results suggest that adding pointing and mapping does not significantly alter SPACE’s ability to distinguish between clinical groups.

**Supplementary Table 7 | Comparison of AUC with and without Pointing and Mapping (PM) measures across CDR groups.** The table reports the difference in AUC (ΔAUC), DeLong test *p*-values, and Bonferroni-corrected DeLong *p*-values for each comparison. Comparisons involving CDR = 2+ were omitted due to low variability in this subgroup.

| **CDR contrasts** | **AUC with PM** | **AUC No PM** | Δ **AUC** | ***p* Delong** | ***p* Bonferroni** |
| --- | --- | --- | --- | --- | --- |
| 0 vs 0.5 | 0.603 | 0.613 | -0.011 | 0.4568 | 1.0000 |
| 0 vs 1 | 0.964 | 0.945 | 0.019 | 0.1224 | 0.3672 |
| 0.5 vs 1 | 0.952 | 0.914 | 0.038 | 0.1330 | 0.3990 |

#### Supplementary Information G

##### ANCOVAs with age and gender

We conducted ANCOVAs to examine whether CDR stage was associated with performance across SPACE tasks, controlling for age and gender. Prior to each ANCOVA, we tested the assumption of homogeneity of regression slopes. All interaction terms were not significant (ps > .10) and therefore removed for parsimony, except for the Memory task, where the Age × CDR interaction was significant (F_2, 243_ = 12.41, p < .001).

There was a significant main effect of CDR on training time (*F*_(2, 276)_ = 17.90, *p* < 0.001), path integration distance error (*F*_(2, 248)_ = 10.69, *p* < 0.001), pointing error (*F*_(2, 247)_ = 3.93, *p* = 0.021), memory percentage correct (*F*_(2, 246)_ = 13.35, *p* < 0.001), and perspective taking error (*F*_(2, 273)_ = 7.49, *p* < 0.001). No significant main effect of CDR was observed for mapping accuracy (*F*_(2, 246)_ = 1.18, *p* = 0.310). Age was a significant covariate for training (*F*_(1, 276)_ = 9.67, *p* = 0.002), path integration (*F*_(1, 248)_ = 27.02, *p* < 0.001), and perspective taking (*F*_(1, 273)_ = 13.17, *p* < 0.001). No significant main effect of gender was detected for any task, though a small CDR-by-gender interaction was present for mapping accuracy (*F*_(2, 246)_ = 3.22, *p* = 0.042). Collectively, these results show that CDR stage explains variance in multiple SPACE measures even after accounting for age and gender.

**Supplementary Table 8** | Results of ANCOVAs examining the main effect of clinical severity (CDR) on performance across SPACE tasks while controlling for age and gender.

| **Task** | **Effect** | **Sum of Squares** | **df** | **Mean Square** | ***F*** | ***p*** |
| --- | --- | --- | --- | --- | --- | --- |
| Training | CDR | 83,225 | 2 | 41,612 | 17.904 | < .001 |
|  | Age | 22,471 | 1 | 22,471 | 9.668 | .002 |
|  | Gender | 2,222 | 1 | 2,222 | 0.956 | .329 |
|  | CDR ✻ Gender | 4,153 | 2 | 2,077 | 0.893 | .410 |
|  | Residuals | 641,492 | 276 | 2,324 |  |  |
| PI Distance | CDR | 150,912 | 2 | 75,456 | 10.693 | < .001 |
|  | Age | 190,635 | 1 | 190,635 | 27.017 | < .001 |
|  | Gender | 7,588 | 1 | 7,588 | 1.075 | .301 |
|  | CDR ✻ Gender | 3,052 | 2 | 1,526 | 0.216 | .806 |
|  | Residuals | 1,750,000 | 248 | 7,056 |  |  |
| Pointing | CDR | 2,664.00 | 2 | 1,332.00 | 3.934 | .021 |
|  | Age | 136.89 | 1 | 136.89 | 0.404 | .525 |
|  | Gender | 1.74 | 1 | 1.74 | 0.005 | .943 |
|  | CDR ✻ Gender | 430.32 | 2 | 215.16 | 0.635 | .531 |
|  | Residuals | 83,641.38 | 247 | 338.63 |  |  |
| Mapping | CDR | 0.2145 | 2 | 0.1072 | 1.175 | .310 |
|  | Age | 0.0866 | 1 | 0.0866 | 0.949 | .331 |
|  | Gender | 0.0123 | 1 | 0.0123 | 0.135 | .714 |
|  | CDR ✻ Gender | 0.5881 | 2 | 0.2941 | 3.223 | .042 |
|  | Residuals | 22.4473 | 246 | 0.0912 |  |  |
| Memory Correct | CDR | 12,270.0 | 2 | 6,135.0 | 13.351 | < .001 |
|  | Age | 85.8 | 1 | 85.8 | 0.187 | .666 |
|  | Gender | 158.0 | 1 | 158.0 | 0.344 | .558 |
|  | CDR ✻ Gender | 1,783.4 | 2 | 891.7 | 1.941 | .146 |
|  | Residuals | 113,042.0 | 246 | 459.5 |  |  |
| Perspective taking | CDR | 5,636 | 2 | 2,818.2 | 7.489 | < .001 |
|  | Age | 4,955 | 1 | 4,954.5 | 13.166 | < .001 |
|  | Gender | 401 | 1 | 401.3 | 1.066 | .303 |
|  | CDR ✻ Gender | 101 | 2 | 50.5 | 0.134 | .875 |
|  | Residuals | 102,733 | 273 | 376.3 |  |  |

##### G.2: ROC analysis including pointing performance

Given that the pointing task was significant in the ANCOVA, we evaluated ROC classification models incorporating SPACE variables (i.e. training, path integration and perspective taking) together with the pointing task, while excluding participants with CDR ≥ 2. The memory task was omitted because of ceiling effects and an imbalanced distribution across CDR levels. Models comparing CDR 0 vs CDR 1 and CDR 0.5 vs CDR 1 showed excellent discrimination (AUC = 0.94, 95% CI: 0.90–0.98; AUC = 0.91, 95% CI: 0.85–0.97, respectively). Performance was more modest for CDR 0 vs CDR 0.5 (AUC = 0.60, 95% CI: 0.52–0.68). Notably, the classification performance was equivalent to that of models that did not include pointing, indicating that adding this metric did not significantly alter diagnostic accuracy.

**Supplementary Table 9 |** ROC analysis for models incorporating SPACE variables, including the pointing task for models excluding CDR 2+. The Memory task was excluded because of ceiling effects and an imbalanced distribution of data across CDR levels.

| **Outcome** | **AUC** | **CI low** | **CI high** | **Cutoff** | **Sens** | **Spec** | **N** |
| --- | --- | --- | --- | --- | --- | --- | --- |
| CDR 0 vs 0.5 | 0.597 | 0.520 | 0.675 | 0.472 | 0.232 | 0.953 | 232 |
| CDR 0 vs 1 | 0.942 | 0.904 | 0.979 | 0.117 | 0.941 | 0.860 | 167 |
| CDR 0.5 vs 1 | 0.909 | 0.850 | 0.968 | 0.112 | 1.000 | 0.744 | 99 |

#### Supplementary Information H

##### Impact of CDR on SPACE after adjusting for critical variables

We conducted robust regressions including anxiety, stress, depression, sleep hours, tablet experience, and self-rated navigation ability (SBSOD) as covariates, in addition to age, gender, and education (Supplementary Table 10). Tablet experience was a significant predictor of training time (*p* = .029), and sleep hours predicted path integration (*p* = .016) and mapping (*p* = .039) performance. Importantly, CDR level remained a significant predictor of training, path integration, and perspective taking performance after accounting for all covariates. When CDR emerged as a significant predictor, we performed post-hoc Tukey-adjusted pairwise comparisons to identify the specific clinical contrasts (Supplementary Table 11).

**Supplementary Table 10 |** Robust linear regression models including depression, anxiety, chronic stress, sleep hours, and prior tablet experience as covariates across SPACE tasks.

|  | **Training** | | | **Path integration** | | | **Pointing** | | | **Mapping** | | | **Perspective taking** | | |
| --- | --- | --- | --- | --- | --- | --- | --- | --- | --- | --- | --- | --- | --- | --- | --- |
| **Predictor** | **Est** | **CI** | ***p*** | **Est** | **CI** | ***p*** | **Est** | **CI** | ***p*** | **Est** | **CI** | ***p*** | **Est** | **CI** | ***p*** |
| (Intercept) | 226.90 | 134.41 – 319.39 | <0.001 | 61.23 | -107.60 – 230.07 | 0.476 | 84.60 | 50.50 – 118.70 | <0.001 | 0.58 | -0.03 – 1.19 | 0.061 | 19.04 | -25.34 – 63.42 | 0.399 |
| CDRglobal [0.5] | 10.40 | -1.83 – 22.62 | 0.095 | 3.09 | -15.75 – 21.93 | 0.747 | 0.61 | -4.66 – 5.88 | 0.820 | 0.01 | -0.08 – 0.10 | 0.819 | -1.45 | -6.34 – 3.44 | 0.561 |
| CDRglobal [1] | 33.19 | 9.65 – 56.73 | 0.006 | 62.69 | 21.35 – 104.04 | 0.003 | 9.40 | -0.51 – 19.30 | 0.063 | 0.12 | -0.02 – 0.27 | 0.088 | 10.19 | 2.86 – 17.53 | 0.007 |
| Age | 0.79 | -0.19 – 1.78 | 0.115 | 2.59 | 1.28 – 3.89 | <0.001 | -0.01 | -0.40 – 0.39 | 0.974 | -0.00 | -0.01 – 0.00 | 0.419 | 0.52 | 0.06 – 0.98 | 0.027 |
| Gender [1] | 7.98 | -3.42 – 19.39 | 0.169 | 22.99 | 5.53 – 40.46 | 0.010 | -0.36 | -5.32 – 4.59 | 0.886 | 0.08 | -0.00 – 0.17 | 0.059 | 0.72 | -4.21 – 5.65 | 0.775 |
| education4levels [1] | -7.96 | -45.19 – 29.28 | 0.674 | -80.90 | -209.86 – 48.05 | 0.218 | 9.86 | -5.24 – 24.96 | 0.200 | -0.14 | -0.39 – 0.12 | 0.292 | -10.27 | -28.49 – 7.94 | 0.268 |
| education4levels [2] | -28.27 | -60.11 – 3.57 | 0.082 | -92.58 | -220.03 – 34.86 | 0.154 | 5.87 | -8.23 – 19.98 | 0.413 | -0.04 | -0.28 – 0.21 | 0.759 | -9.94 | -27.06 – 7.18 | 0.254 |
| education4levels [3] | -30.53 | -62.93 – 1.87 | 0.065 | -82.33 | -211.03 – 46.37 | 0.209 | 4.77 | -9.54 – 19.08 | 0.512 | 0.01 | -0.23 – 0.26 | 0.925 | -13.42 | -30.69 – 3.85 | 0.127 |
| education4levels [4] | -19.57 | -52.92 – 13.78 | 0.249 | -94.64 | -223.15 – 33.86 | 0.148 | 2.56 | -11.80 – 16.93 | 0.726 | 0.01 | -0.24 – 0.26 | 0.957 | -18.85 | -35.99 – -1.71 | 0.031 |
| Anxiety | -1.88 | -6.51 – 2.75 | 0.425 | -2.10 | -9.01 – 4.80 | 0.549 | 0.60 | -1.63 – 2.84 | 0.596 | 0.02 | -0.01 – 0.05 | 0.208 | -0.92 | -2.84 – 1.00 | 0.347 |
| ChrStress | 1.14 | -3.20 – 5.49 | 0.605 | 5.00 | -1.31 – 11.31 | 0.120 | 0.10 | -1.98 – 2.18 | 0.925 | -0.03 | -0.06 – 0.00 | 0.070 | 1.35 | -0.68 – 3.38 | 0.191 |
| Depression | -0.64 | -5.83 – 4.54 | 0.807 | -0.53 | -9.40 – 8.34 | 0.907 | -0.97 | -3.14 – 1.20 | 0.378 | 0.01 | -0.02 – 0.05 | 0.389 | 0.51 | -1.72 – 2.73 | 0.653 |
| SleepHrs | 1.33 | -2.44 – 5.10 | 0.488 | 4.79 | 0.91 – 8.67 | 0.016 | -0.26 | -1.66 – 1.13 | 0.710 | 0.02 | 0.00 – 0.04 | 0.039 | -1.06 | -3.00 – 0.88 | 0.281 |
| ExpTablet [1] | -9.34 | -28.79 – 10.10 | 0.345 | 4.29 | -26.59 – 35.18 | 0.784 | -2.33 | -9.96 – 5.30 | 0.548 | -0.00 | -0.13 – 0.12 | 0.959 | -2.96 | -10.61 – 4.70 | 0.448 |
| ExpTablet [2] | -20.91 | -39.68 – -2.14 | 0.029 | -7.09 | -37.47 – 23.29 | 0.646 | -6.25 | -14.02 – 1.52 | 0.114 | 0.07 | -0.06 – 0.20 | 0.274 | -6.93 | -14.80 – 0.94 | 0.084 |
| SBSOD | 2.50 | -2.80 – 7.81 | 0.354 | -3.13 | -10.41 – 4.14 | 0.397 | -1.30 | -3.74 – 1.14 | 0.294 | -0.02 | -0.06 – 0.02 | 0.383 | 2.48 | -0.04 – 4.99 | 0.054 |
| Observations | 283 | | | 255 | | | 254 | | | 253 | | | 280 | | |
| R2 / R2 adjusted | 0.225 / 0.182 | | | 0.307 / 0.264 | | | 0.073 / 0.014 | | | 0.086 / 0.028 | | | 0.211 / 0.167 | | |

##### H.2: Post hoc Tukey-adjusted pairwise comparisons for significant predictor from previous regression model.

**Table 11 |** **Robust pairwise post hoc contrasts (Tukey-adjusted) for the effect of clinical diagnosis (CDR) across SPACE task outcomes.** Models were fitted with robust linear regression (*lmrob*) controlling for age, gender, education, depression, anxiety, chronic stress, sleep hours, tablet experience, and SBSOD. Significance codes: *** p < .001, ** p < .01, * p < .05, ns = not significant.

| **Outcome** | **Contrast (CDR)** | **Estimate** | **SE** | **z** | **p** |  |
| --- | --- | --- | --- | --- | --- | --- |
| **Training Time** | 0.5 – 0 | 10.395 | 6.211 | 1.674 | 0.204 | *ns* |
|  | 1 – 0 | 33.192 | 11.957 | 2.776 | 0.014 | *** |
|  | 1 – 0.5 | 22.797 | 12.688 | 1.797 | 0.161 | *ns* |
| **Path Integration Distance** | 0.5 – 0 | 3.090 | 9.564 | 0.323 | 0.940 | *ns* |
|  | 1 – 0 | 62.695 | 20.989 | 2.987 | 0.007 | **** |
|  | 1 – 0.5 | 59.605 | 23.266 | 2.562 | 0.025 | *** |
| **Perspective Error** | 0.5 – 0 | -1.448 | 2.489 | -0.582 | 0.827 | *ns* |
|  | 1 – 0 | 10.189 | 3.736 | 2.727 | 0.017 | *** |
|  | 1 – 0.5 | 11.637 | 3.679 | 3.163 | 0.004 | **** |

#### Supplementary Information I

##### SPACE Performance Between APOE ε4 Carriers and Non-Carriers

We conducted exploratory analyses to examine whether APOE ε4 carrier status was associated with performance in SPACE. None of the tasks significantly differentiated carriers from non-carriers. Because genotype data were available only for a smaller, imbalanced subsample (n = 96), these results were excluded from the main analysis. No significant differences were observed for training time (W = 828, p = .46, 95% CI = –34.89 to 14.53, median difference = –10.43), path integration (W = 524, p = .60, 95% CI = –52.21 to 35.98, median difference = –7.87), pointing (W = 557, p= .89, 95% CI = –10.62 to 9.37, median difference = –0.75), mapping (W = 593, p = .71, 95% CI = –0.12 to 0.22, median difference = 0.03), or perspective taking (W = 869, p = .77, 95% CI = –11.19 to 8.35, median difference = –1.55).

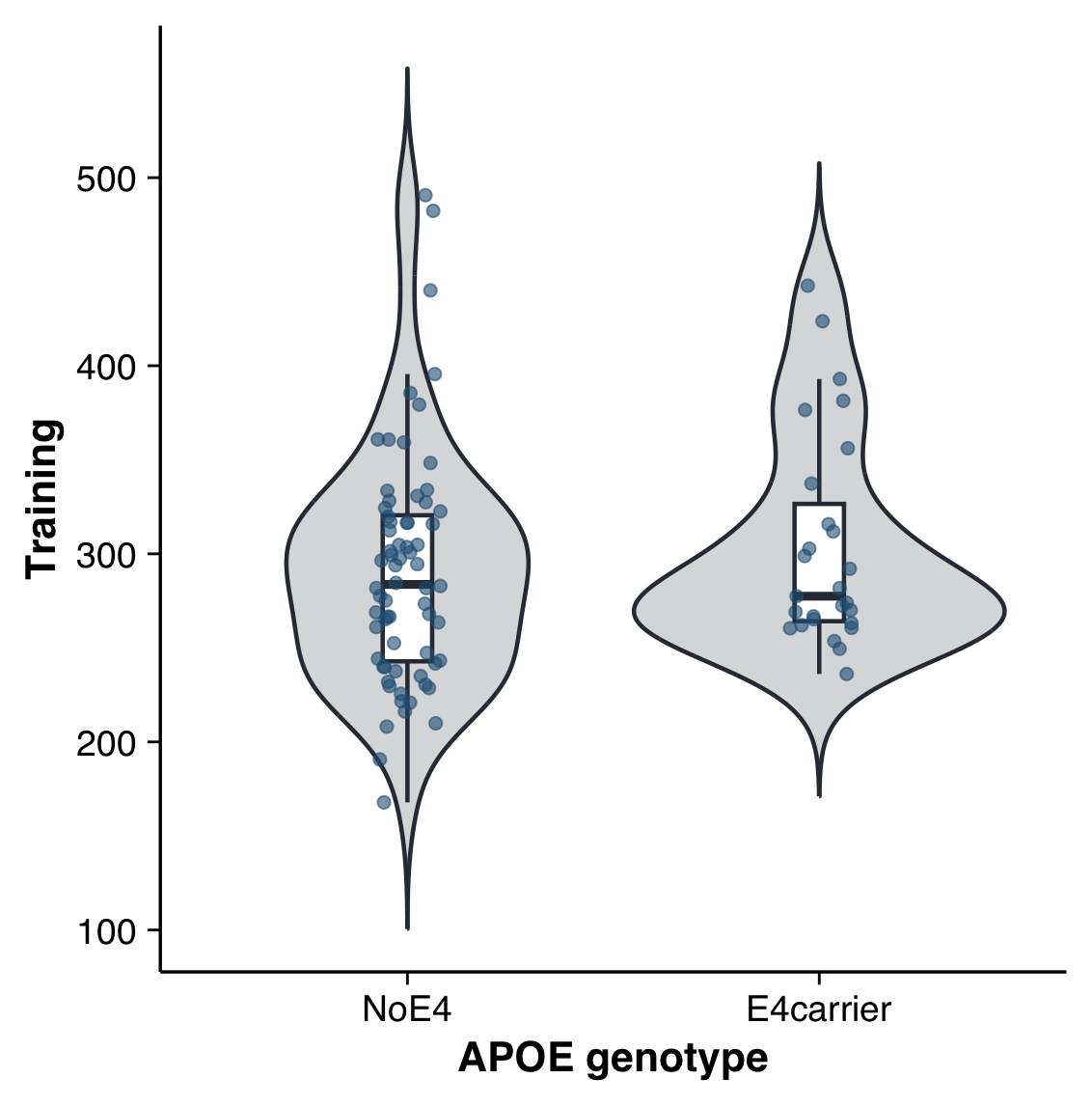

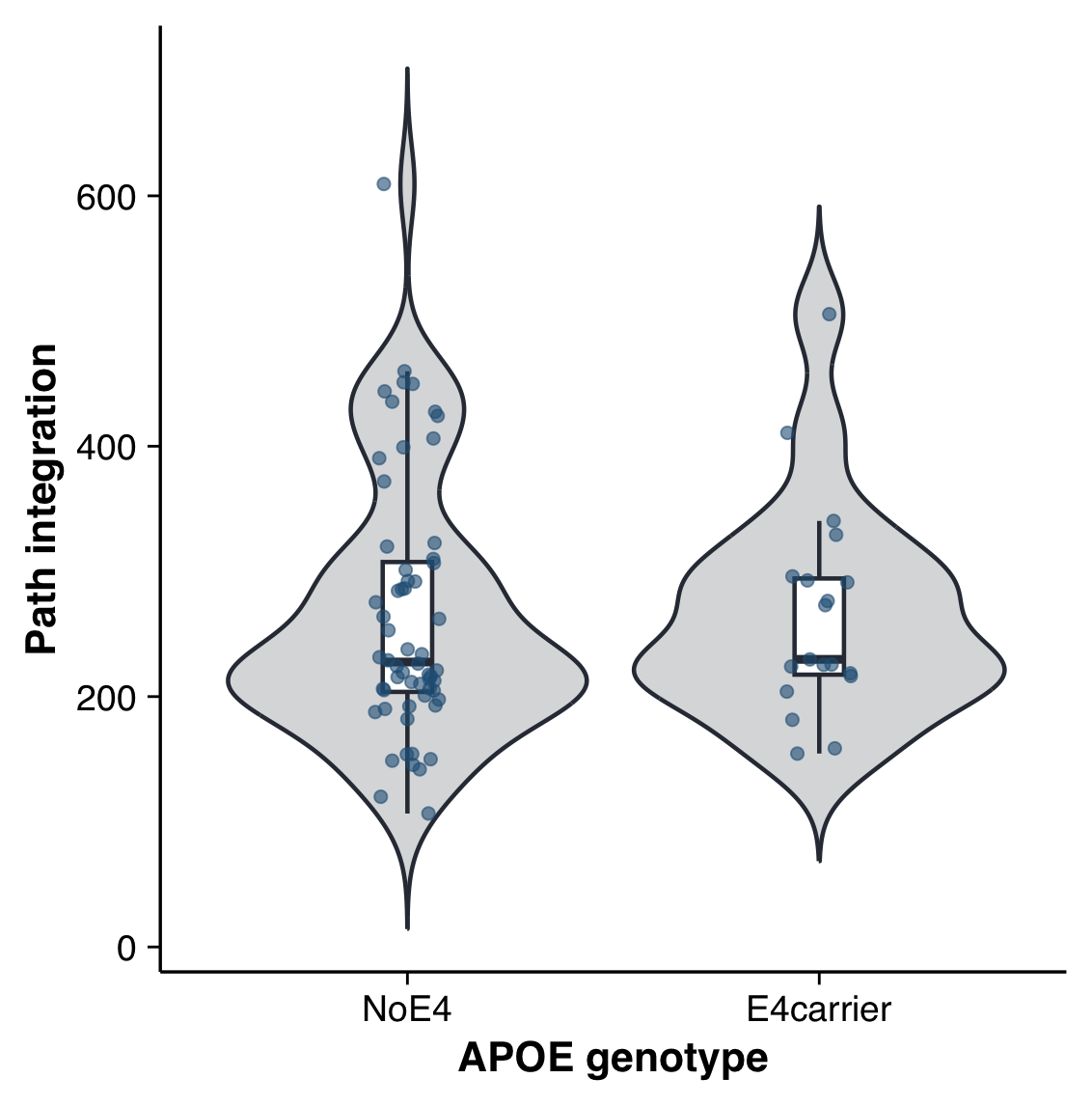

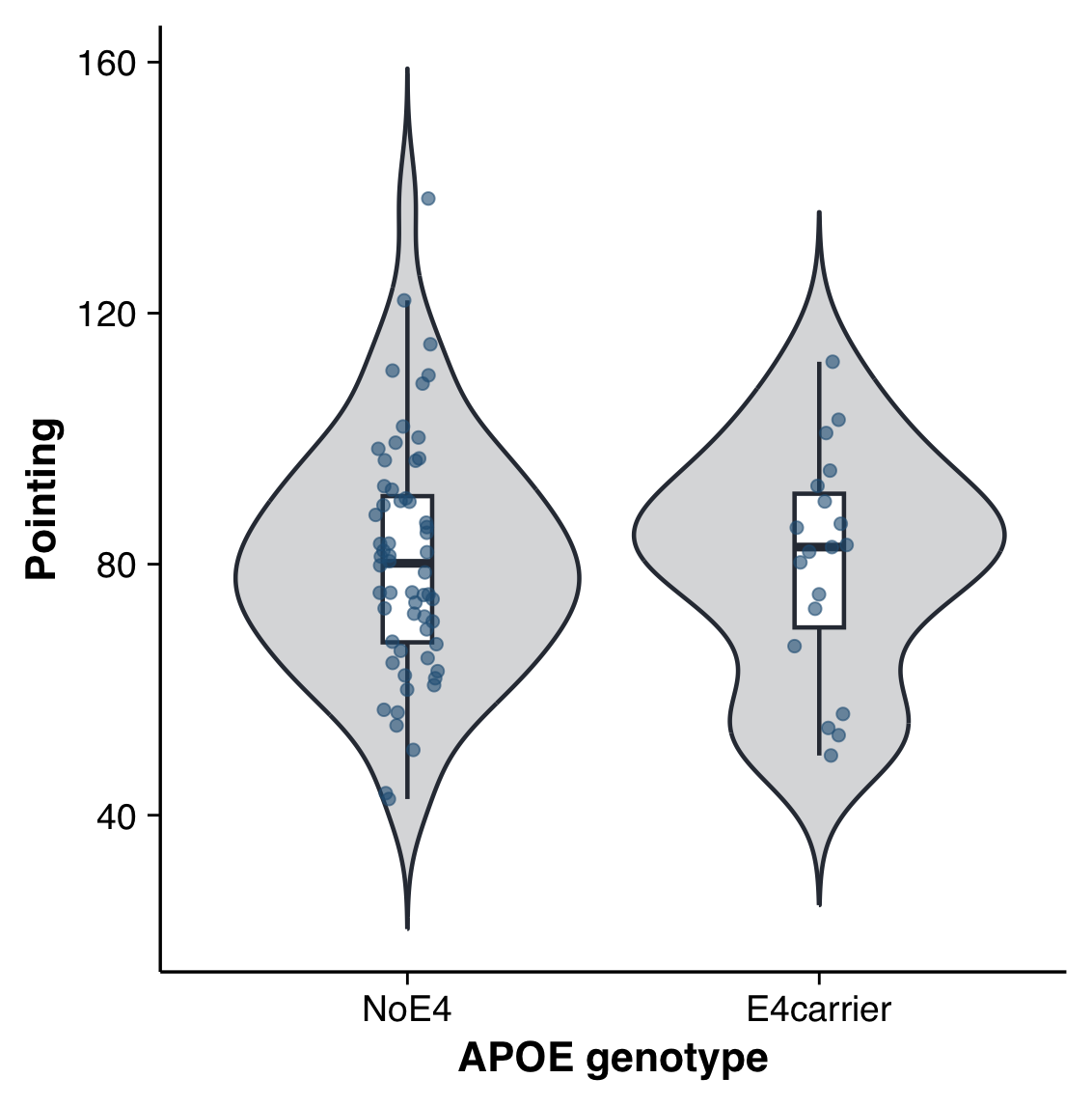

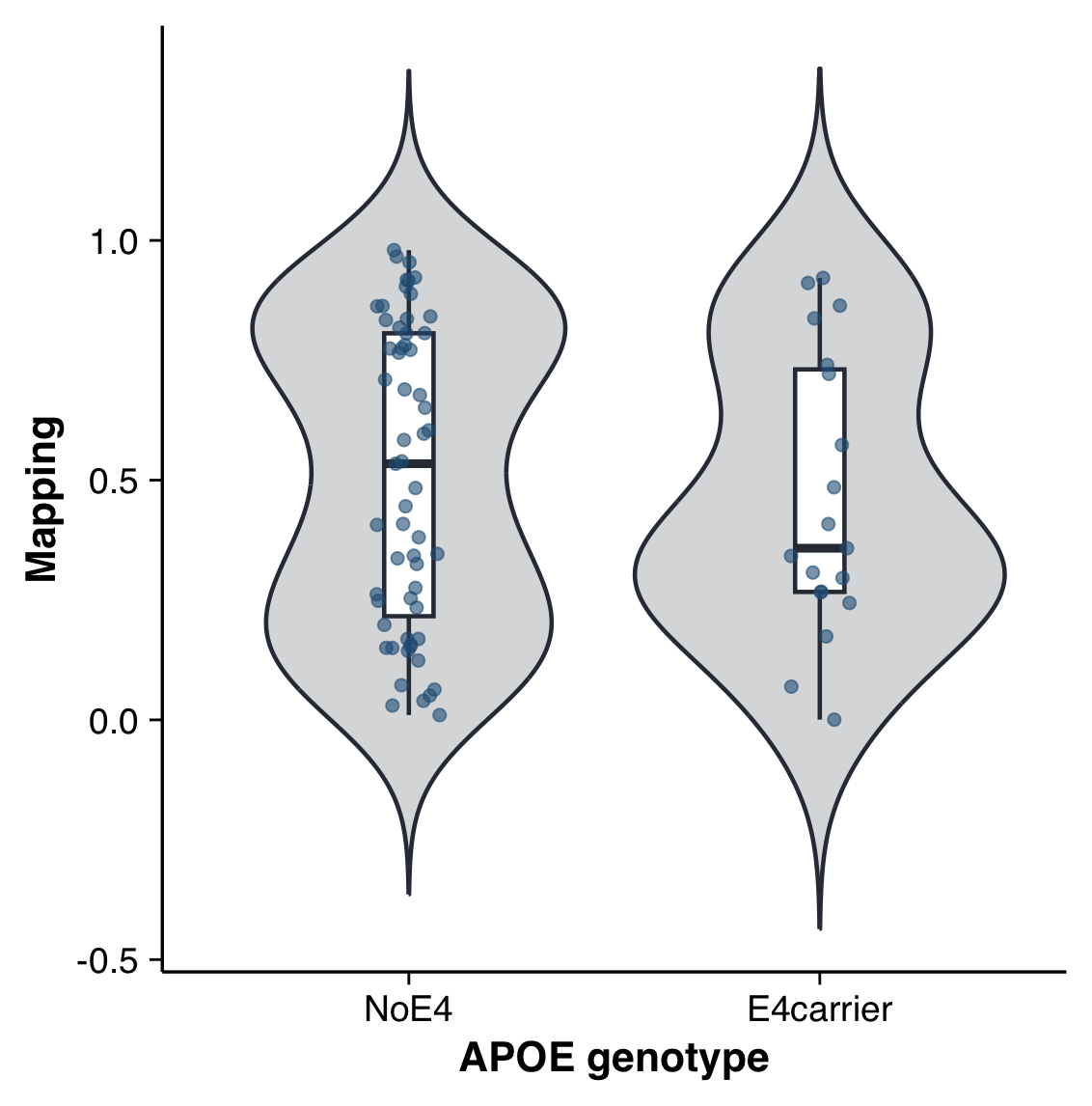

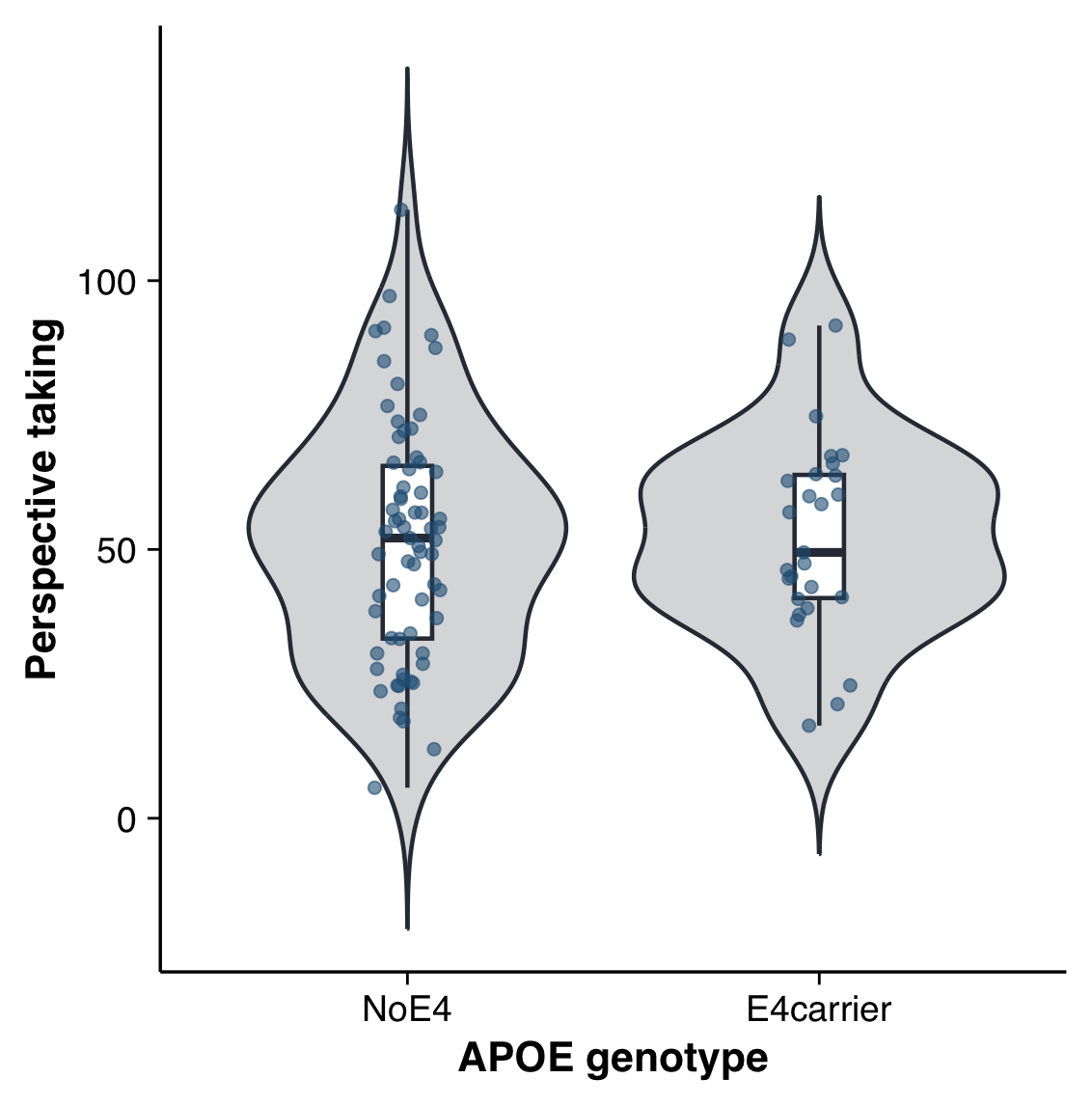

**Supplementary Figure 2 |** ***Performance across the tasks in SPACE for ε4 carriers vs non-carriers.*** Violin plots for the performance in the tasks in SPACE between APOE4 carriers and non-carriers, with embedded box plots and individual data points. No significant group differences were detected.

#### Supplementary Information J

##### Other measures

###### The Vascular Dementia Battery (VDB)

The VDB is a validated, detailed neuropsychological test battery administered to all subjects by trained research psychologists. The test battery assessed seven cognitive domains, including executive function, attention, language, visuomotor speed, visuoconstruction, visual memory and verbal memory.

The VDB is composed of a series of tests that evaluate different cognitive domains: i. Executive Function: Frontal Assessment Battery. ii. Attention: Digit Span, Visual Memory Span and Auditory Detection. iii. Language: Language: Modified Boston Naming Test and Verbal Fluency. iv. Visuomotor Speed: Symbol Digit Modality Test, Maze Task and Digit Cancellation. v. Visuoconstruction: Weschler Memory Scale – Revised (WMS-R) Visual Reproduction Copy task, Clock Drawing and Weschler Adult Intelligence Scale – Revised (WAIS-R) subtest of Block Design. vi. Visual Memory: Picture Recall & Recognition Tasks, and WMS-R Visual Reproduction Recall & Recognition Task. vii. Verbal Memory: Word List Recall & Recognition Tasks and Story Recall.

###### Neuropsychological assessment

A comprehensive neuropsychological battery was administered by the certified psychometrists and used to assess multiple cognitive domains of participants. It consisted of the Quick Dementia Rating System (QDRS), Montreal Cognitive Assessment (MoCA), maze task, Digit Cancellation Test (DCT), Trail Making Test (TMT), and dual-task test.

QDRS: The QDRS is a 10-item questionnaire completed by an informant that rates a patient’s cognitive functioning in 10 domains (i.e., memory and recall, orientation, problem-solving, activities outside the home, functioning at home, personal hygiene, behaviour and personality changes, language and communication, mood, and attention). Scores range from 0 to 30, with higher scores indicating worse cognitive impairment.

MoCA: The MoCA is a widely used screening tool for detecting cognitive impairment by evaluating visuospatial abilities, executive function, language, attention, short-term memory, and orientation. Scores range from 0 to 30, with lower scores indicating worse cognitive impairment. A score of 25 or below is indicative of MCI, while a score of 18 or below suggests dementia.

Maze Task: The maze task was used to assess the cognitive abilities related to spatial and visual perception. During the test, participants are presented with a maze on a piece of paper and are asked to find their way out by drawing a line from the entrance to the exit as quickly as possible. The outcome variable is the duration needed to complete the task, with a faster time indicating better performance.

Digit Cancellation Test (DCT): The DCT was developed to measure attention. In this task, participants are required to cross out target digits printed on a page mixed with other numbers within 45 seconds. The final score is calculated as the subtraction of the number of incorrectly cancelled digits from the total number of correctly cancelled digits. The higher the final score, the better the performance.

Trail Making Test (TMT): The TMT was used to measure attention, visual screening ability, and processing speed. The test consists of two parts, A and B. In part A, participants are asked to connect the circles with numbers in ascending order. In part B, participants are required to connect the circles by switching between numbers and letters in consecutive order (e.g., 1, A, 2, B). The time to completion in seconds was reported separately for each part of the TMT test and used for scoring. The shorter the time, the better the performance.

Dual-task: The dual-task test was used to assess the ability to perform two tasks concurrently. This paper-and-pencil dual-task consisted of performing digit recall and tracking tasks separately and then simultaneously. First, each participant underwent a digit span assessment to determine their maximum digit span capacity. This was followed by two trials that involved both digit recall and a tracking task to familiarise the participants with the dual-task test. For the digit recall task, a list of numbers was read to the participants, and they were required to verbally repeat the numbers in the exact order in which they were read. For the tracking task, participants had to trace a predefined route through the paper with a pencil, joining all the circles as quickly as possible. Each task was restricted to 1.5 minutes. After familiarisation, the dual-task test was administered with the same time limit, and subsequently, participants were required to complete both tasks simultaneously. Proportional performance in both tasks combined was calculated as the final performance score.

Animal fluency test: The animal fluency task consisted of naming as many animals as possible within a one-minute time frame. The final score reflects the total number of animals correctly named.

###### Fluid biomarkers and APOE status

A total of 15 mL of blood was collected through venesection into ethylenediaminetetraacetic acid (EDTA) tubes. The tubes were centrifuged at 2000 crf for 1 minute, 4°C, and the upper plasma layers were extracted and stored at -80°C until analysed. Plasma biomarker levels were measured on Simoa HD-X instruments (Quanterix, Billerica, MA, USA) at the National University of Singapore (NUS), using the Neuro 3-plex assay (for the detection of Aβ40, Aβ42, t-tau) and p-tau217, all purchased from Quanterix (Billerica, MA, USA).

Apolipoprotein E (APOE) genotype was assessed by the polymerase chain reaction followed by restriction fragment length polymorphism (PCR-RFLP) analysis using the blood samples collected from the patients^63^. The presence of at least one APOE ε4 allele was regarded as APOE ε4-positive.

###### Gait assessment

Gait function was assessed using wearable inertial measurement units (WT901BLECL BLE 5.0, WITMOTION, Shenzhen, China), which integrated high-precision gyroscopes, accelerometers, and geomagnetic field sensors. Here, five synchronised sensors were placed on the different body parts (one on the trunk, one on each foot, and one on each hand) and connected to an iPad via Bluetooth. Gait parameters were measured under the single-task (baseline) and dual-task conditions. In the baseline condition, participants were asked to walk along the walkway without talking. The dual-task condition consisted of walking while reciting out loud serial subtractions of three, starting from 400. Under both single- and dual-task conditions, participants walked along the same path in the same corridor at their self-selected walking speed for three minutes.
